# Continuum of Maternal Healthcare and Neonatal Mortality in Sub-Saharan Africa

**DOI:** 10.64898/2026.08.17.26360589

**Authors:** John Senanu, Precious Fafali Dotse, Ellen Osei Ephson

## Abstract

**Background:** Neonatal mortality remains a significant public health challenge in Sub-Saharan Africa (SSA). The continuum of maternal care (COC), spanning antenatal care (ANC), skilled birth attendance (SBA), and postnatal care (PNC) represents an integrated pathway to improving neonatal survival. Multi-country evidence on how adherence to this full continuum affects neonatal outcomes across SSA is limited.

**Objectives:** This study aimed to: (1) construct a composite COC indicator and describe its sociodemographic distribution; (2) estimate its association with neonatal mortality; (3) conduct a counterfactual analysis; and (4) examine cross-country heterogeneity in the COC effect on neonatal mortality.

**Methods:** Pooled Demographic and Health Survey (DHS) data from 35 SSA countries (2010– 2026) were analyzed (N = 867,984 live births). A binary CoC indicator (coc1 = 1 if mother received ≥4 ANC visits, skilled birth attendance, and PNC within 48 hours) was constructed. Survey-weighted logistic regression adjusted for wealth, education, residence, parity, maternal age, child sex, child age, and country. Counterfactual predictive margins and a COC × country interaction model were estimated in Stata 18.

**Results:** Only 13.47% of mothers met the full COC threshold. COC completion was higher among wealthier, urban, more educated, and lower-parity women. After adjustment, CoC receipt was associated with significantly lower odds of neonatal death (aOR = 0.638, 95% CI: 0.577– 0.706, p < 0.001). Counterfactual analysis showed the predicted neonatal mortality probability would fall from 3.15% (no CoC) to 2.04% (full CoC), an absolute risk reduction of 1.11 percentage points. Cross-country interaction terms were largely non-significant; only Namibia reached significance (p = 0.036).

**Conclusion:** Completion of the full continuum of maternal care is independently associated with reduced neonatal mortality across SSA. Equity-focused policies should prioritize integrated service delivery for rural, poor, and less-educated women.

## Introduction

Neonatal mortality, death within the first 28 days of life, represents one of the most critical challenges in global child health. Sub-Saharan Africa (SSA) bears a disproportionate burden, accounting for approximately 40% of all global neonatal deaths despite comprising a smaller fraction of global births (United Nations Inter-agency Group for Child Mortality Estimation [UN IGME], 2023). In 2022, the neonatal mortality rate in SSA averaged 27 deaths per 1,000 live births, far exceeding the Sustainable Development Goal (SDG) 3.2 target of 12 deaths per 1,000 live births by 2030 (World Health Organization [WHO], 2023).

The leading causes of neonatal death; preterm birth complications, intrapartum-related events, and neonatal sepsis are largely preventable through timely and integrated maternal and newborn care (Liu et al., 2016). The continuum of care (COC) framework posits that linking ANC, skilled birth attendance, and early postnatal care in an uninterrupted and sequential manner substantially reduces adverse neonatal outcomes (Kerber et al., 2007). While each component independently reduces neonatal mortality risk, the synergistic benefits of the full continuum have been less systematically evaluated at the multi-country level in SSA.

Considerable heterogeneity exists in COC uptake across SSA. Socioeconomic inequalities, geographic barriers, and health system weaknesses contribute to fragmented care, with many women accessing one or two components but failing to complete the full continuum (Amouzou et al., 2014; Delvaux et al., 2017). This study used data from the Demographic and Health Surveys of 35 SSA countries to: (1) describe COC distribution across sociodemographic groups; (2) estimate the adjusted association between COC and neonatal mortality; (3) conduct counterfactual analyses; and (4) examine cross-country heterogeneity in the COC and neonatal mortality association.

## Methods

### Study Design and Data Source

A cross-sectional, multi-country pooled analysis was conducted using publicly available Demographic and Health Survey(DHS) data from 35 Sub Saharan Afican countries (survey years 2010–2026). The DHS Program conducts nationally representative household surveys using standardized questionnaires, multi-stage stratified cluster sampling, and probability-proportional-to-size selection. Women’s (IR), births (BR), children’s (KR), and household (HR) files were merged. Ethical clearance was not required as DHS data are fully anonymized and publicly available.

### Study Population and Variables

The analytical sample comprised all live births within five years preceding each survey (N = 867,984). The primary outcome was neonatal mortality (death within 28 days of birth). The composite CoC indicator (coc1 = 1) required satisfaction of all three criteria: ≥4 ANC visits, skilled birth attendance, and postnatal care within 48 hours. Covariates included wealth index (v190), child sex (b4), country (country1), maternal education (v106), continuous maternal age (v012), child age at survey (ageatsurveydate), place of residence (v025rr), and parity (v201).

### Statistical Analysis

Descriptive statistics used chi-square tests with column percentages. Survey-weighted logistic regression (Model 1) estimated adjusted odds ratios (aOR) using Stata’s svy prefix. Counterfactual predictive margins were estimated with the margins command. Cross-country heterogeneity was assessed using a coc1 × country1 interaction model (Model 2). All analyses used linearized standard errors to account for complex survey design. Statistical significance was set at p < 0.05.

## Results

Wealth index. CoC receipt increased monotonically with wealth quintile: only 16.19% of poorest-quintile mothers completed the CoC compared to 24.08% of richest-quintile mothers (chi-squared(4) = 18,000, p < 0.001). Conversely, 27.95% of non-CoC births were in the poorest quintile, compared to just 16.19% in the CoC group. Figure 1 illustrates this gradient clearly.

**Figure 1.**
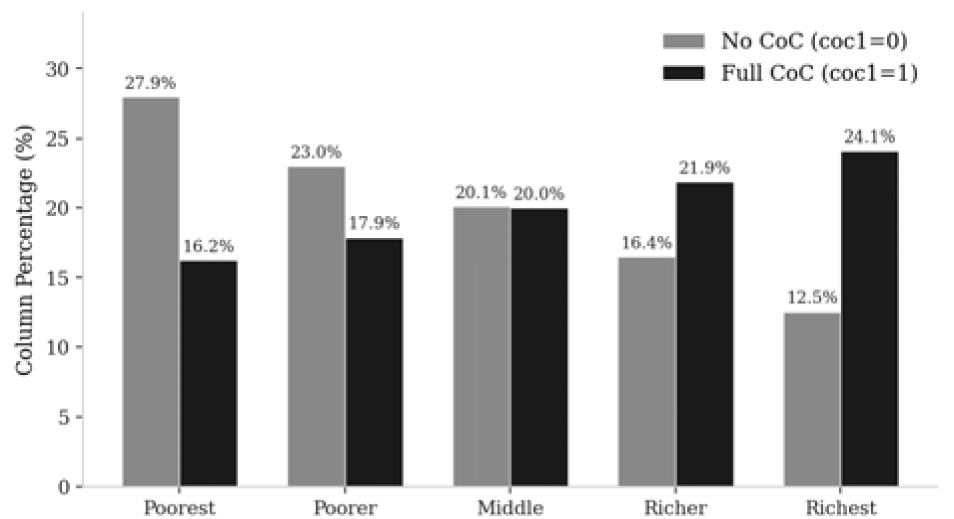
Continuum of Care Completion by Household Wealth Quintile. Column percentages within each CoC group. Pearson chi-squared(4) = 18,000, p < 0.001. COC = Continuum of Care.

Education. Figure 2 illustrates the education gradient in CoC completion. Among non-CoC mothers, 45.39% had no education versus only 25.25% in the CoC group. Secondary education was markedly higher in the CoC group (37.66% vs. 20.01% in non-CoC), and higher education increased from 2.35% (non-CoC) to 8.45% (CoC). Residence: Urban mothers were more likely to complete the CoC (44.16% in CoC vs. 28.02% in non-CoC; chi-squared(1) = 13,000, p < 0.001). Parity: CoC prevalence declined with increasing parity; first-time mothers accounted for 25.82% of CoC births versus only 12.73% of non-CoC births (chi-squared(3) = 18,000, p < 0.001).

**Figure 2.**
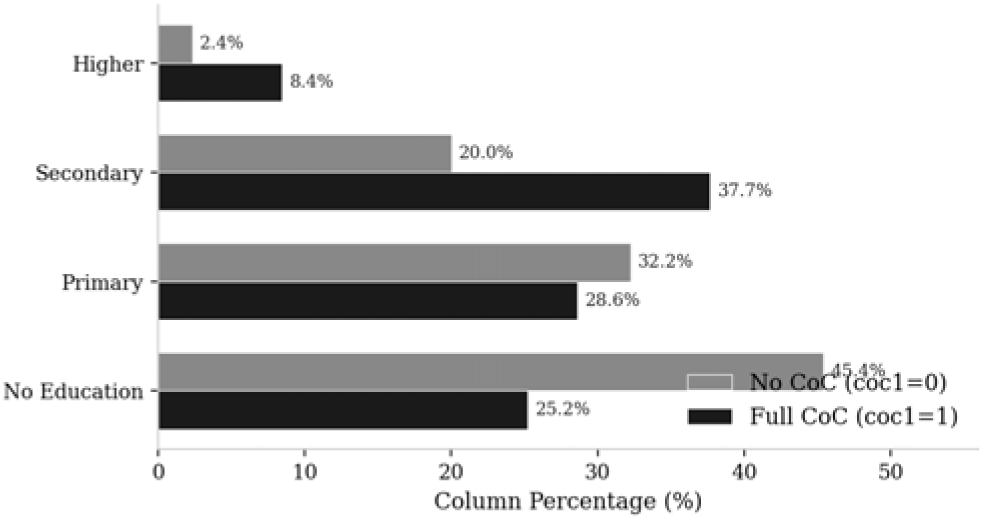
Continuum of Care Completion by Maternal Educational Level. *Note. Column percentages within each CoC group. Pearson chi-squared(5) = 36,000, p < 0.001*.

Figure 3 visualizes the joint distributions by parity and residence. The decline in CoC completion with increasing parity is evident in panel (a), while the urban advantage in CoC uptake is clear in panel (b). Country-level variation: Substantial between-country variation in CoC prevalence was observed (chi-squared(34) = 30,000, p < 0.001), examined in detail in Figure 6.

**Figure 3.**
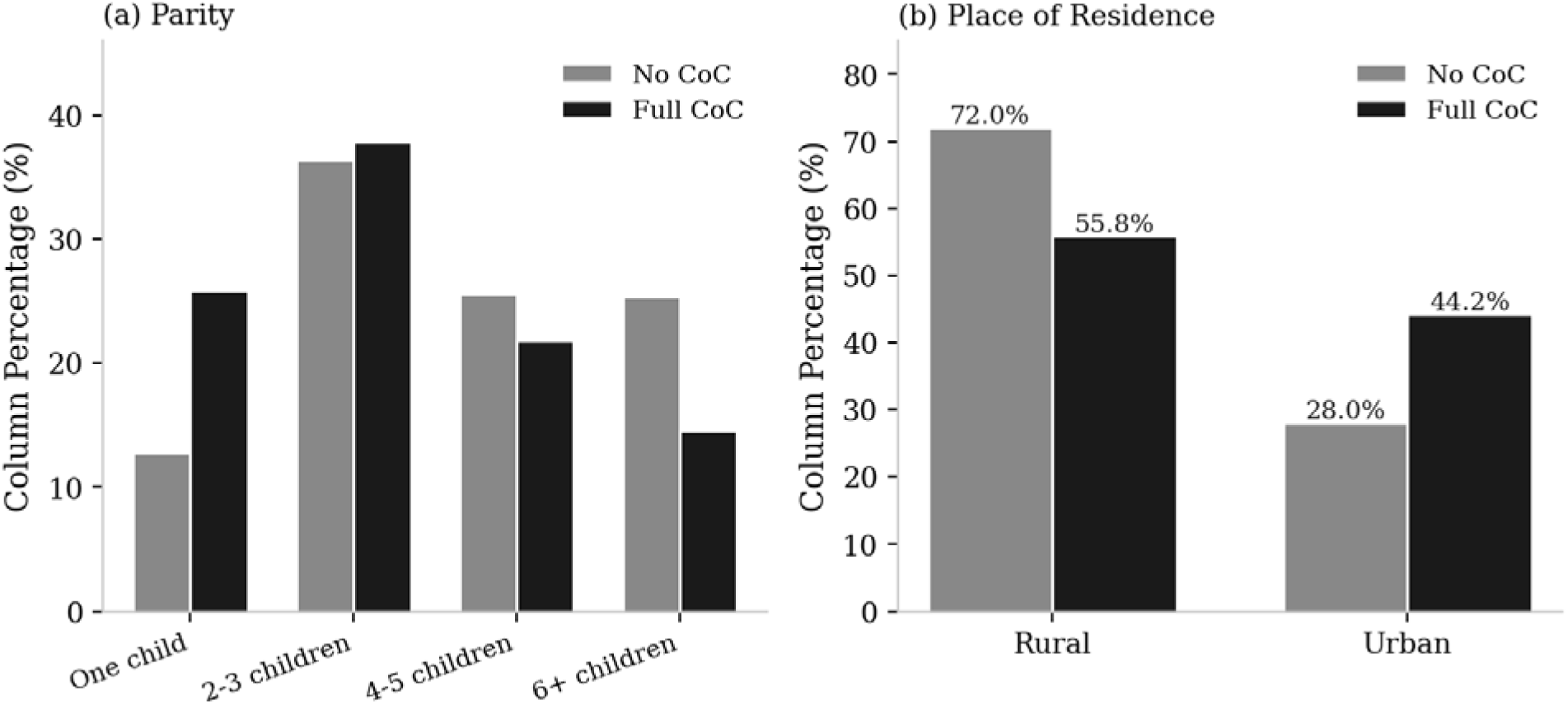
Continuum of Care Completion by Parity and Place of Residence. *Note. Pan el (a) shows CoC distribution by parity; panel (b) by urban/rural residence. Column percentages within each CoC group*.

### Association Between Continuum of Care and Neonatal Mortality (Model 1)

Table 2 presents adjusted odds ratios from the survey-weighted logistic regression (N = 865,362; F(9, 37186) = 78.66, p < 0.001).

**Table 1.** Bivariate Distribution of Continuum of Care (COC1) by Sociodemographic.

*Characteristics*
| Variable | CoC = 0 (n, %) | CoC = 1 (n, %) | p-value |
| --- | --- | --- | --- |
| **Wealth Index** |  |  |  |
| Poorest | 209,778 (27.95%) | 18,908 (16.19%) | <0.001 |
| Poorer | 172,622 (23.00%) | 20,854 (17.85%) |  |
| Middle | 150,785 (20.09%) | 23,360 (20.00%) |  |
| Richer | 123,467 (16.45%) | 25,556 (21.88%) |  |
| Richest | 93,982 (12.52%) | 28,132 (24.08%) |  |
| **Sex of Child** |  |  |  |
| Male | 379,831 (50.57%) | 59,763 (51.11%) | 0.001 |
| Female | 371,214 (49.43%) | 57,176 (48.89%) |  |
| **Maternal Education** |  |  |  |
| No education | 340,913 (45.39%) | 29,526 (25.25%) | <0.001 |
| Primary | 242,188 (32.25%) | 33,487 (28.64%) |  |
| Secondary | 150,274 (20.01%) | 44,041 (37.66%) |  |
| Higher | 17,622 (2.35%) | 9,880 (8.45%) |  |
| **Place of Residence** |  |  |  |
| Rural | 540,589 (71.98%) | 65,299 (55.84%) | <0.001 |
| Urban | 210,456 (28.02%) | 51,640 (44.16%) |  |
| **Parity** |  |  |  |
| One child | 95,573 (12.73%) | 30,191 (25.82%) | <0.001 |
| 2-3 children | 272,902 (36.34%) | 44,302 (37.88%) |  |
| 4-5 children | 192,090 (25.58%) | 25,477 (21.79%) |  |
| 6+ children | 190,480 (25.36%) | 16,969 (14.51%) |  |
| **Maternal Age Group** |  |  |  |
| 15-19 years | 43,741 (5.82%) | 8,269 (7.07%) | <0.001 |
| 20-24 years | 168,389 (22.42%) | 26,311 (22.50%) |  |
| 25-29 years | 202,250 (26.93%) | 29,935 (25.60%) |  |
| 30-34 years | 156,326 (20.81%) | 24,766 (21.18%) |  |
| 35-39 years | 110,967 (14.78%) | 17,642 (15.09%) |  |
| 40-44 years | 52,559 (7.00%) | 7,858 (6.72%) |  |
| 45-49 years | 16,813 (2.24%) | 2,158 (1.85%) |  |
| **Child Age at Survey** |  |  |  |
| 0 months | 6,270 (0.83%) | 1,490 (1.27%) | <0.001 |
| 1-11 months | 135,092 (17.99%) | 34,118 (29.18%) |  |
| 12-23 months | 137,830 (18.35%) | 35,309 (30.19%) |  |
| 24-35 months | 143,398 (19.09%) | 26,477 (22.64%) |  |
| 36-47 months | 165,032 (21.97%) | 11,424 (9.77%) |  |
| 48-59 months | 163,423 (21.76%) | 8,121 (6.94%) |  |

CoC = Continuum of Care (coc1). Percentages are column percentages. All chi-square tests $p < 0.001$ unless noted.

**Table 2.** Survey-Weighted Logistic Regression: Adjusted Odds Ratios for Neonatal Mortality (Model 1)

| Variable | aOR | SE | t | p | 95% CI |
| --- | --- | --- | --- | --- | --- |
| Full CoC – coc1=1 | 0.638** | 0.033 | -8.73 | <.001 | [0.577, 0.706] |
| Wealth index (v190) | 1.039** | 0.015 | 2.63 | .008 | [1.010, 1.070] |
| Child sex – Female (b4) | 0.740** | 0.023 | -9.69 | <.001 | [0.696, 0.786] |
| Country (c_ountry1) | 1.008** | 0.001 | 5.37 | <.001 | [1.005, 1.011] |
| Maternal education (v106) | 1.050* | 0.020 | 2.54 | .011 | [1.011, 1.091] |
| Maternal age – continuous (v012) | 0.952** | 0.004 | -11.99 | <.001 | [0.944, 0.960] |
| Child age at survey (continuous) | 0.998* | 0.001 | -2.24 | .025 | [0.997, 1.000] |
| Urban residence (v025rr) | 1.010 | 0.043 | 0.23 | .822 | [0.928, 1.099] |
| Parity (v201) | 1.192** | 0.012 | 17.11 | <.001 | [1.168, 1.216] |

*Note.* aOR = adjusted odds ratio; SE = linearized standard error; CI = confidence interval. Reference: coc1 = 0. \* $p < 0.05$ ; \*\* $p < 0.01$ .

Continuum of care. Figure 4 presents the forest plot of all model covariates. Mothers completing the full CoC had 36.2% lower odds of neonatal death (aOR = 0.638, 95% CI: 0.577– 0.706, p < 0.001) compared to non-CoC mothers—the strongest protective association in the model. Parity was the strongest risk factor (aOR = 1.192, 95% CI: 1.168–1.216, p < 0.001). Female children had lower odds of neonatal death than males (aOR = 0.740, 95% CI: 0.696– 0.786, p < 0.001). Older maternal age was protective (aOR = 0.952/year, p < 0.001). Urban residence was not independently significant (aOR = 1.010, p = 0.822) after adjusting for other factors.

**Figure 4.**
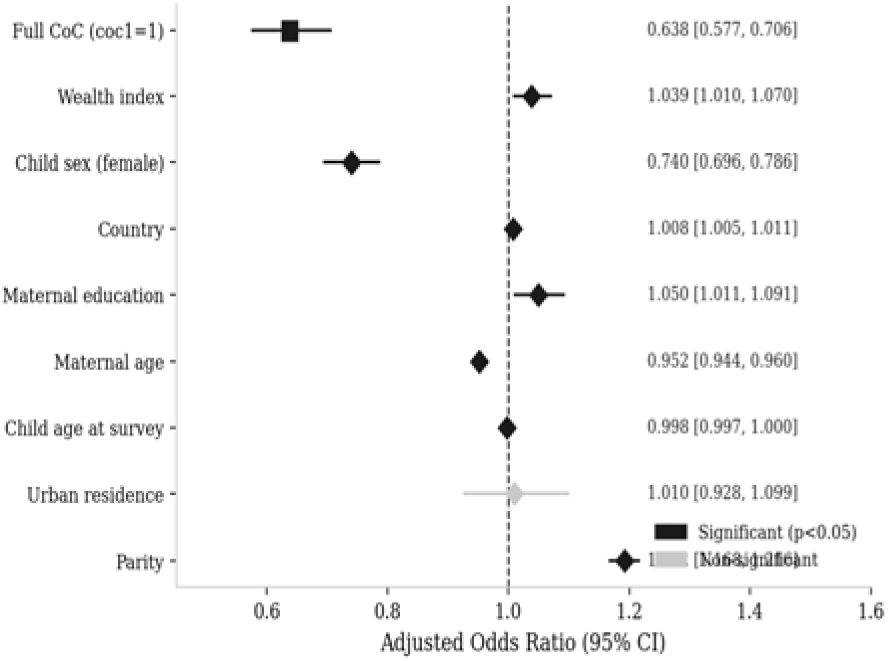
Forest Plot of Adjusted Odds Ratios for Neonatal Mortality (Model 1) *Note. Square markers = statistically significant (p < 0.05; dark blue); diamond markers = non-significant (grey). Horizontal lines denote 95% confidence intervals. Dashed vertical line at OR = 1.0*.

### Counterfactual Analysis: Predictive Margins and Average Marginal Effects

Table 3 presents the estimated predictive margins and average marginal effect of CoC on neonatal mortality probability.

**Table 3.** Predictive Margins and Average Marginal Effects of Continuum of Care on Neonatal Mortality.

| Scenario | Predicted Prob. | SE | t | p-value | 95% CI |
| --- | --- | --- | --- | --- | --- |
| **Predictive Margins** |  |  |  |  |  |
| No CoC (coc1 = 0) | 0.0315 | 0.0005 | 62.04 | <0.001 | [0.0305, 0.0325] |
| Full CoC (coc1 = 1) | 0.0204 | 0.0010 | 19.79 | <0.001 | [0.0184, 0.0224] |
| **Average Marginal Effect** |  |  |  |  |  |
| CoC dy/dx | -0.0111 | 0.0011 | -10.47 | <0.001 | [-0.0132, -0.0091] |

*Note. Margins estimated with other covariates held at observed values (N = 864,824). AME = average marginal effect.*

Figure 5 visually contrasts the counterfactual scenarios. Without CoC, the predicted probability of neonatal death was 3.15% (95% CI: 3.05–3.25%). With universal CoC adherence, this would fall to 2.04% (95% CI: 1.84–2.24%)—an absolute risk reduction of 1.11 percentage points (dy/dx = -0.0111, SE = 0.0011, t = -10.47, p < 0.001). This represents a relative risk reduction of approximately 35.2%. Applied across the 867,984 births in this pooled sample, universal CoC adherence could potentially avert approximately 9,635 neonatal deaths.

**Figure 5.**
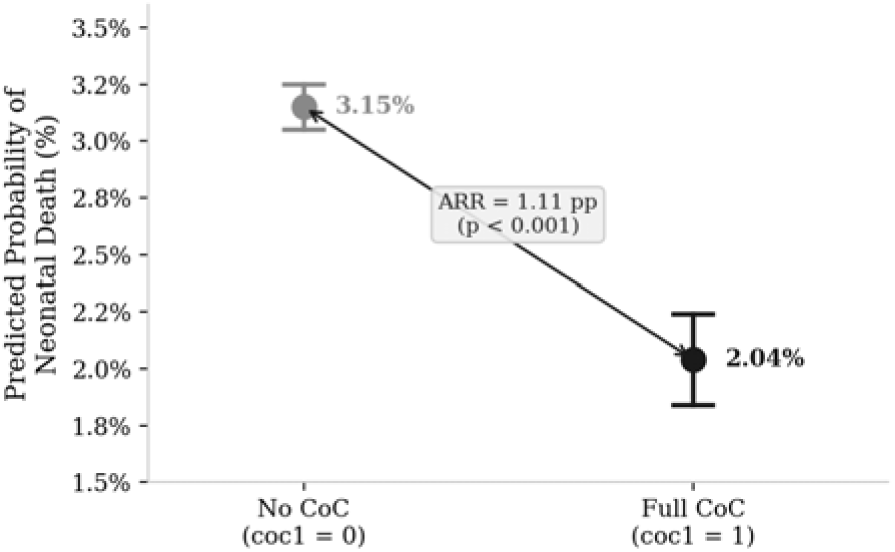
Counterfactual Predictive Margins for Neonatal Mortality by Continuum of Care Status. *Note. Points = predicted probability of neonatal death; error bars = 95% confidence intervals. ARR = absolute risk reduction. The two-headed arrow illustrates the 1.11 percentage-point reduction associated with full CoC completion*.

### Cross-Country Heterogeneity in the CoC Effect (Model 2)

Table 4 presents selected country-specific CoC × country interaction terms from Model 2 (F(76, 37119) = 17.33, p < 0.001).

**Table 4.** Cross-Country Heterogeneity: Selected CoC x Country Interaction Terms (Model 2)

| Country | Interaction OR | SE | t | p | 95% CI |
| --- | --- | --- | --- | --- | --- |
| CoC main effect (Angola ref.) | 0.669 | 0.186 | -1.45 | .147 | [0.388, 1.152] |
| Namibia x CoC | 0.414* | 0.174 | -2.10 | .036 | [0.182, 0.942] |
| Rwanda x CoC | 0.470 | 0.195 | -1.82 | .069 | [0.209, 1.061] |
| Ghana x CoC | 0.538 | 0.192 | -1.74 | .082 | [0.268, 1.081] |
| Zimbabwe x CoC | 0.534 | 0.201 | -1.66 | .096 | [0.255, 1.118] |
| South Africa x CoC | 0.564 | 0.222 | -1.45 | .146 | [0.260, 1.221] |
| Nigeria x CoC | 0.980 | 0.283 | -0.07 | .944 | [0.557, 1.724] |
| Senegal x CoC | 0.889 | 0.270 | -0.39 | .698 | [0.491, 1.611] |
| Malawi x CoC | 0.742 | 0.226 | -0.98 | .327 | [0.409, 1.348] |
| Kenya x CoC | 0.793 | 0.256 | -0.72 | .472 | [0.422, 1.491] |
| Burkina Faso x CoC | 0.707 | 0.235 | -1.04 | .298 | [0.369, 1.357] |

Note. Interaction OR = odds ratio for CoC × country interaction; reference country = Angola. Only Namibia was statistically significant ( $p = 0.036$ ). \* $p < 0.05$ .

Figure 6 displays the country-level distribution of CoC completion proportions within the CoC group. Nigeria (14.29%), Malawi (7.22%), and Zambia (5.51%) had the highest representation, reflecting both large sample sizes and relatively higher coverage. Chad (0.55%), Democratic Republic of Congo (0.93%), and Guinea (0.89%) had the lowest representations, consistent with known health system constraints in these contexts. Despite this between-country variation in baseline CoC and neonatal mortality rates, the interaction model indicated that the protective effect of CoC on neonatal mortality was broadly consistent across countries (Table 4). Only Namibia showed a statistically significant interaction term (interaction OR = 0.414, 95% CI: 0.182–0.942, p = 0.036), indicating a stronger CoC protective effect there relative to Angola. Rwanda (p = 0.069) and Ghana (p = 0.082) showed borderline non-significant stronger CoC effects.

**Figure 6.**
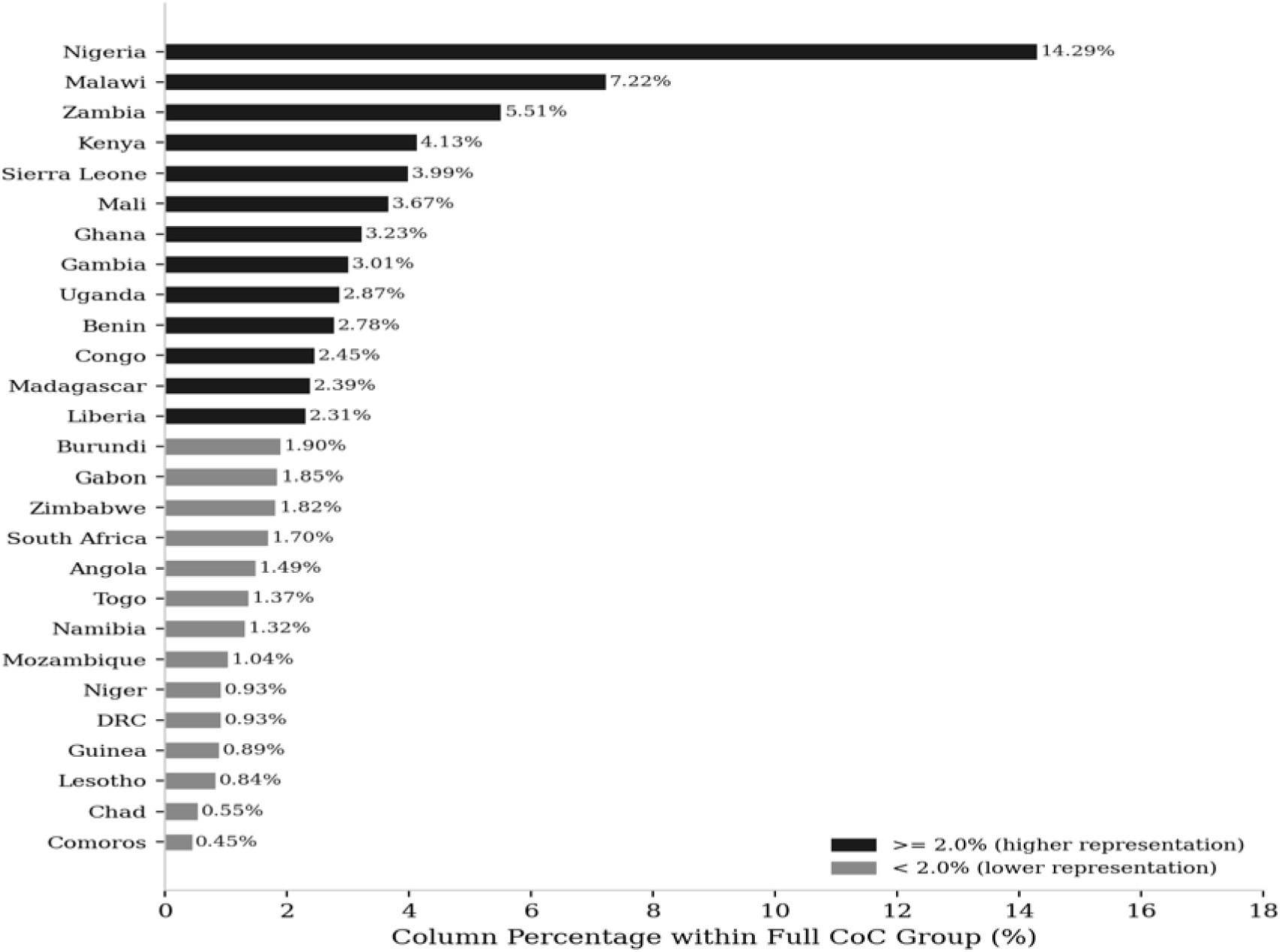
Country-Level Distribution of Continuum of Care Completion in Sub-Saharan Africa. *Note. Bars show column percentage within the full CoC group (coc1 = 1) by country. Dark blue = countries with* ≥*2.0% share; grey = <2.0%. Countries ordered by ascending CoC representation*.

## Discussion

The analysis of DHS data from 35 Sub Saharan African countries provides robust, population-level evidence that completion of the full continuum of maternal care is significantly associated with reduced neonatal mortality. The adjusted odds ratio of 0.638 (95% CI: 0.577– 0.706) indicates that CoC-compliant mothers had approximately 36% lower odds of neonatal death after accounting for wealth, education, residence, parity, maternal age, child characteristics, and country. These findings are consistent with prior evidence from individual-country studies and meta-analyses highlighting the cumulative benefits of integrated maternal care (Bhutta et al., 2014; Kerber et al., 2007).

The counterfactual analysis (Figure 5) quantified that universal COC adherence could reduce the predicted neonatal mortality probability from 3.15% to 2.04%, an absolute risk reduction of 1.11 percentage points and relative reduction of 35.2%. Applied across the pooled sample, this could avert approximately 9,635 neonatal deaths, underscoring the public health significance of closing the COC gap.

The wealth and education gradients shown in Figures 1 and 2 reveal persistent and deep inequalities in COC uptake. Poorer, rural, less-educated, and higher-parity women remain least likely to complete the full continuum (Figure 3), mirroring long-established health service utilization inequities in SSA (Fotso et al., 2009). With only 13.47% of all mothers meeting the full COC threshold, the gap between current coverage and what is needed to achieve SDG 3.2 neonatal mortality targets is enormous.

The country-level distribution of COC coverage (Figure 6) and the cross-country interaction model (Table 4) together reveal that while baseline neonatal mortality rates and COC coverage vary substantially across SSA countries, the COC protective effect itself is broadly homogeneous. Only Namibia achieved a statistically significant interaction term, suggesting country-specific factors such as stronger health system integration or postnatal care quality may amplify the CoC benefit there. This overall consistency of effect across diverse settings strengthens the case for region-wide COC promotion policies.

The forest plot (Figure 4) contextualizes the CoC effect among all covariates. Parity remained the strongest independent risk factor for neonatal death (aOR = 1.192), consistent with evidence that grand multiparous women face elevated obstetric risks (Mgaya et al., 2013). The female sex survival advantage (aOR = 0.740) is biologically well-documented (Drevenstedt et al., 2008). The non-significance of urban residence after full adjustment confirms that the urban– rural differential in neonatal mortality is largely explained by socioeconomic and educational factors rather than residence per se.

### Limitations

This study has several limitations. First, the cross-sectional DHS design precludes causal inference. Second, COC was based on self-reported data subject to recall bias, particularly for births 3–5 years before the survey evidenced by declining COC prevalence in older birth cohorts. Third, the composite COC indicator captures service receipt but not quality. Fourth, pooling surveys from 2010–2026 may introduce period effects, though country fixed effects partially address this. Fifth, unmeasured community-level supply-side factors may still confound results.

### Policy Implications

The findings carry several important implications for maternal and neonatal health policy in Sub-Saharan Africa.

1. Integrated service delivery. Health systems should move from vertical programs toward integrated care packages ensuring seamless ANC intrapartum postnatal linkages. Task-shifting to community health workers, mobile outreach, and COC tracking tools such as maternity passports can bridge service gaps.
2. Targeting vulnerable groups. Interventions must prioritize poor, rural, uneducated, and high-parity women with the lowest COC completion rates. Conditional cash transfers, social protection programs, and community-based postnatal care have demonstrated effectiveness.
3. Postnatal care scale-up. As postnatal care within 48 hours is often the most commonly missed CoC component, dedicated investments in PNC awareness, facility capacity, and community PNC delivery are essential.
4. Country-specific strategies. Countries with very low COC coverage DRC, Chad, Angola require context-specific strategies addressing both demand-side barriers and supply-side constraints.
5. Monitoring and accountability. National health information systems should include COC completion as a routine monitoring indicator tracked jointly with SDG 3.2 neonatal mortality targets.

## Conclusion

This large-scale pooled DHS analysis from 35 Sub Saharan African countries demonstrates that completion of the full continuum of maternal care is independently and significantly associated with a 36% reduction in neonatal mortality odds. The counterfactual analysis (Figure 5) quantifies a population-level absolute risk reduction of 1.11 percentage points. With only 13.47% of mothers meeting the full CoC threshold, and the deepest deficits among the poorest, most rural, least educated, and highest-parity women (Figures 1–3), equity-focused integrated care delivery is urgently needed. The broadly consistent COC protective effect across SSA countries (Figure 6; Table 4) supports region-wide policy commitments to integrated maternal-neonatal care. Accelerating COC coverage is critical to achieving SDG 3.2 neonatal mortality targets across Sub-Saharan Africa.

## Data Availability

All data produced in the present study are available upon reasonable request to the authors

## Notes

### Competing Interest Statement

The authors have declared no competing interest.

### Author Declarations

Existing public datasets were used.

## References

Amouzou, A., Habi, O., & Bensaid, K. (2014). Reduction in child mortality in Niger: A Countdown to 2015 country case study. The Lancet, 383(9930), 1756–1762. 10.1016/S0140-6736(14)60621-9

Bhutta, Z. A., Das, J. K., Bahl, R., Lawn, J. E., Salam, R. A., Paul, V. K., Sankar, M. J., Blencowe, H., Rizvi, A., Chou, V. B., & Walker, N. (2014). Can available interventions end preventable deaths in mothers, newborn babies, and stillbirths, and at what cost? The Lancet, 384(9940), 347–370. 10.1016/S0140-6736(14)60792-3

Delvaux, T., Konan, J. P., Ake-Tano, O., Gohou-Kouassi, V., Bosso, P. E., Buve, A., & Ronsmans, C. (2017). Quality of antenatal and delivery care before and after the implementation of a prevention of mother-to-child HIV transmission programme in Cote d’Ivoire. Tropical Medicine and International Health, 12(8), 1018–1028. 10.1111/j.1365-3156.2007.01882.x

Drevenstedt, G. L., Crimmins, E. M., Vasunilashorn, S., & Finch, C. E. (2008). The rise and fall of excess male infant mortality. Proceedings of the National Academy of Sciences, 105(13), 5016–5021. 10.1073/pnas.0800221105

Fotso, J. C., Ezeh, A., Madise, N., Ziraba, A., & Ogollah, R. (2009). What does access to maternal care mean among the urban poor? Maternal and Child Health Journal, 13(1), 130–137. 10.1007/s10995-008-0326-4

Kerber, K. J., de Graft-Johnson, J. E., Bhutta, Z. A., Okong, P., Starrs, A., & Lawn, J. E. (2007). Continuum of care for maternal, newborn, and child health: From slogan to service delivery. The Lancet, 370(9595), 1358–1369. 10.1016/S0140-6736(07)61578-5

Liu, L., Oza, S., Hogan, D., Chu, Y., Perin, J., Zhu, J., Lawn, J. E., Cousens, S., Mathers, C., & Black, R. E. (2016). Global, regional, and national causes of under-5 mortality in 2000–15. The Lancet, 388(10063), 3027–3035. 10.1016/S0140-6736(16)31593-8

Mgaya, A. H., Massawe, S. N., Kidanto, H. L., & Mgaya, H. N. (2013). Grand multiparity: Is it still a risk in pregnancy? BMC Pregnancy and Childbirth, 13(1), Article 241. 10.1186/1471-2393-13-241

StataCorp. (2023). Stata statistical software: Release 18. StataCorp LLC.

United Nations Inter-agency Group for Child Mortality Estimation. (2023). Levels and trends in child mortality: Report 2023. UNICEF.

Victora, C. G., Requejo, J. H., Barros, A. J., Berman, P., Bhutta, Z., Boerma, T., Chopra, M., de Francisco, A., Daelmans, B., Hazel, E., Lawn, J., Maliqi, B., Newby, H., & Bryce, J. (2016). Countdown to 2015: A decade of tracking progress for maternal, newborn, and child survival. The Lancet, 387(10032), 2049–2059. 10.1016/S0140-6736(15)00519-X

World Health Organization. (2023). World health statistics 2023: Monitoring health for the SDGs. WHO Press.

